# Large language models in radiologic numerical tasks: A thorough evaluation and error analysis

**DOI:** 10.1101/2025.10.16.25337607

**Authors:** Ali Nowroozi, Masha Bondarenko, Adrian Serapio, Tician Schnitzler, Sukhmanjit S Brar, Jae Ho Sohn

**Affiliations:** Center for Intelligent Imaging, Department of Radiology and Biomedical Imaging, University of California, San Francisco (UCSF), San Francisco, California; All India Institute of Medical Sciences, Bhopal, India

**Keywords:** Large language models, data extraction, radiology reports, reasoning, mathematics, numbers

## Abstract

**Purpose:** To investigate the performance of LLMs in radiology numerical tasks and perform a comprehensive error analysis.

**Materials and Methods:** We defined six tasks: extracting 1-minimum T-score from DEXA report, 2-maximum common bile duct (CBD) diameter from ultrasound report, and 3-maximum lung nodule size from CT report, and judging 1-presence of a highly hypermetabolic region on a PET report, 2-whether a patient is osteoporotic based on a DEXA report, and 3-whether a patient has a dilated CBD based on an ultrasound report. Reports were extracted from the MIMIC III and our institution’s databases, and the ground truths were extracted manually. The models used were Llama 3.1 8b, DeepSeek R1 distilled Llama 8b, OpenAI o1-mini, and OpenAI GPT-5-mini. We manually reviewed all incorrect outputs and performed a comprehensive error analysis.

**Results:** In extraction tasks, while Llama showed relatively variable results (ranging 86%-98.7%) across tasks, other models performed consistently well (accuracies >95%). In judgement tasks, the lowest accuracies of Llama, DeepSeek, o1-mini, and GPT-5-mini were 62.0%, 91.7%, 91.7%, and 99.0%, respectively, while o1-mini and GPT-5-mini did reach 100% performance in detecting osteoporosis. We found no mathematical errors in the outputs of o1-mini and GPT-5-mini. Answer-only output format significantly reduced performance in Llama and DeepSeek but not in o1-mini or GPT-5-mini.

**Conclusion:** True reasoning models perform consistently well in radiology numerical tasks and show no mathematical errors. Simpler non-true reasoning models may also achieve acceptable performance depending on the task.

## Introduction

Large language models (LLMs) are being increasingly utilized for various clinical and research tasks in the medical domain, including clinical diagnoses, management suggestions, and report refinements (1,2). There is an exponentially growing body of medical research involving LLMs (3), with varying degrees of performance across applications (1,4). Known limitations of these models leading to suboptimal outcomes include hallucinations, limited input size, and susceptibility to changes in instruction prompts (1).

One previously documented weakness of LLMs has been their difficulty with numerical reasoning and calculations (5,6), which has also recently been a focus of attention in medical studies involving LLMs and has been suggested as a source for suboptimal LLM performance (1,7). In a study that tested LLMs in assigning Reporting and Data Systems (RADS) categories, numerical errors were partly responsible for incorrect cases (8). Newer models incorporating iterative ‘reasoning’ capabilities have been shown to perform exceedingly better than older-generation models (9,10) in numerical tasks. However, the use of these agents has been limited by limited API access, high processing costs, large model sizes, and privacy concerns (10–12), and reasoning induction using chain-of-thought prompting in older models might not necessarily lead to better performance (13).

Although these challenges have been iterated in previous studies (1,14) and to some degree explored in healthcare-related scenarios (15,16), there is a paucity of research on the performance of LLMs in clinically relevant radiological numerical tasks. In this study, we aimed to thoroughly assess the performance of both open-weight and proprietary state-of-the-art LLMs in numerical tasks derived from radiological texts. In addition, we comprehensively analyzed the errors that the models made to better understand their behavior and how to approach such tasks in future research work and real-world practices.

## Methods

### Multi-step extraction and judgment tasks

LLMs have been previously shown to be adept at basic retrieval tasks (i.e., extracting information clearly present in the final intended form in the text) (17,18). We extended this task and designed two tasks to more comprehensively evaluate the numerical reasoning abilities of LLMs: 1) multi-step information extraction (extracting numbers involving mathematical operations such as comparison and unit conversion), and 2) clinical number-related judgment. The following clinically relevant report-specific tasks were selected, encompassing a wide variety of inputs and objectives:

Extraction:

1. Extracting the maximum reported nodule measurement from a CT report
2. Extracting the maximum common bile duct (CBD) diameter from an ultrasound report
3. Extracting the minimum T-score from a dual-energy X-ray absorptiometry (DEXA) report.

Judgment:

1. Determining the presence of a highly hypermetabolic region in a positron emission tomography (PET) report, as defined by SUVMax > 5, except in areas with expected normal uptake such as the brain.
2. Determining whether a patient has osteoporosis based on T-scores on a DEXA report.
3. Determining whether a patient has a dilated common bile duct based on an ultrasound report.

The exact prompts and criteria can be found in Table 1. We used simplified criteria to isolate the model’s core numerical reasoning abilities without overshadowing them with nuanced clinical complexities. Furthermore, as our main goal was not maximizing model performance by all means, we did not perform rigorous prompt engineering. Nonetheless, there are a few sections of the prompts that were set by preliminary trial and error: a) we had to mention that the models only consider current values, since that reflects the real-world clinical uses of the tasks and models sometimes did not automatically consider that, and b) we included the term “be as concise as possible” to prevent infinite loops and encourage the models to provide short responses that fit within the set output token limit. All ground truth extraction, evaluation, and error analyses were done by two independent reviewers (AN, MB), and discrepancies were resolved by discussion.

**Table 1.**
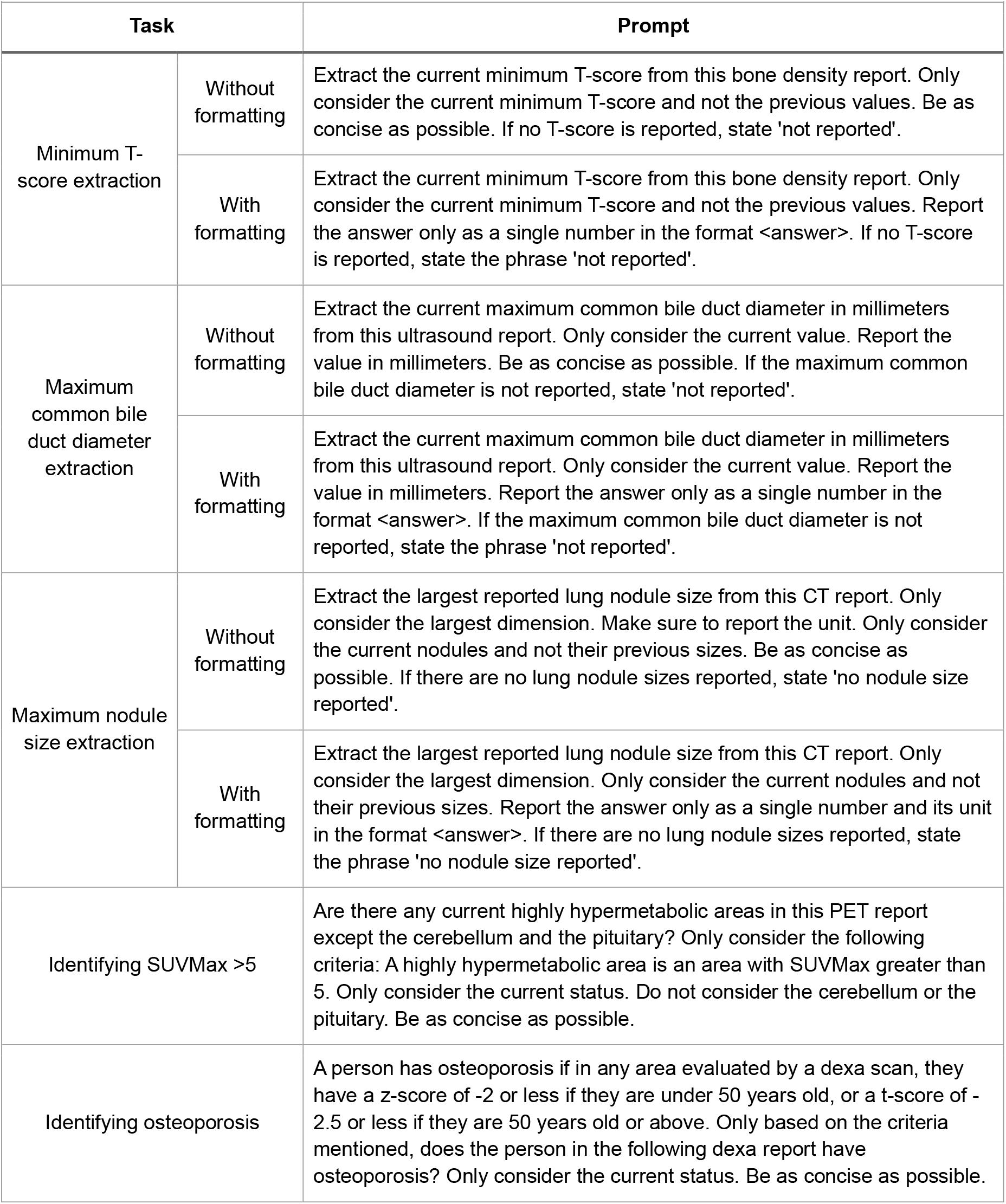
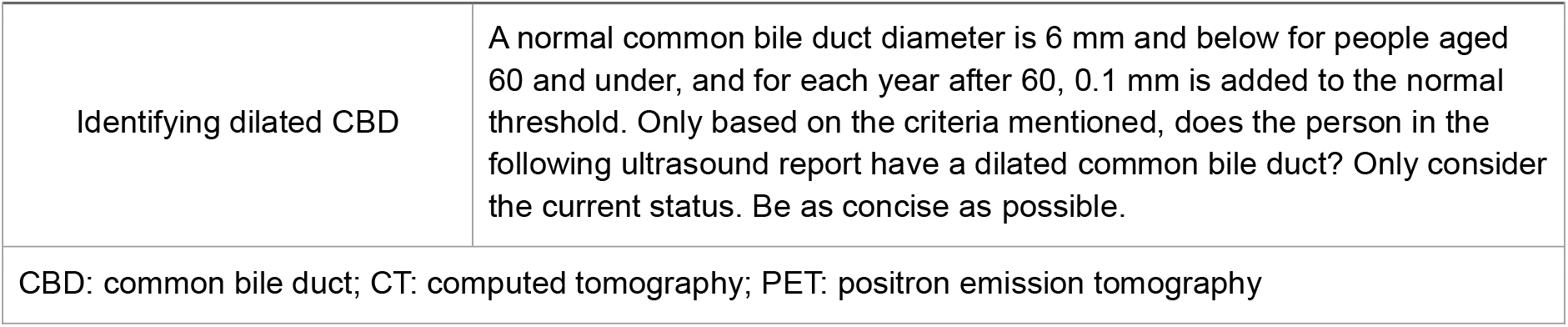
Prompts given to the models.

| Task |  | Prompt |
| --- | --- | --- |
| Minimum T-score extraction | Without formatting | Extract the current minimum T-score from this bone density report. Only consider the current minimum T-score and not the previous values. Be as concise as possible. If no T-score is reported, state 'not reported'. |
|  | With formatting | Extract the current minimum T-score from this bone density report. Only consider the current minimum T-score and not the previous values. Report the answer only as a single number in the format <answer>. If no T-score is reported, state the phrase 'not reported'. |
| Maximum common bile duct diameter extraction | Without formatting | Extract the current maximum common bile duct diameter in millimeters from this ultrasound report. Only consider the current value. Report the value in millimeters. Be as concise as possible. If the maximum common bile duct diameter is not reported, state 'not reported'. |
|  | With formatting | Extract the current maximum common bile duct diameter in millimeters from this ultrasound report. Only consider the current value. Report the value in millimeters. Report the answer only as a single number in the format <answer>. If the maximum common bile duct diameter is not reported, state the phrase 'not reported'. |
| Maximum nodule size extraction | Without formatting | Extract the largest reported lung nodule size from this CT report. Only consider the largest dimension. Make sure to report the unit. Only consider the current nodules and not their previous sizes. Be as concise as possible. If there are no lung nodule sizes reported, state 'no nodule size reported'. |
|  | With formatting | Extract the largest reported lung nodule size from this CT report. Only consider the largest dimension. Only consider the current nodules and not their previous sizes. Report the answer only as a single number and its unit in the format <answer>. If there are no lung nodule sizes reported, state the phrase 'no nodule size reported'. |
| Identifying SUVMax >5 |  | Are there any current highly hypermetabolic areas in this PET report except the cerebellum and the pituitary? Only consider the following criteria: A highly hypermetabolic area is an area with SUVMax greater than 5. Only consider the current status. Do not consider the cerebellum or the pituitary. Be as concise as possible. |
| Identifying osteoporosis |  | A person has osteoporosis if in any area evaluated by a dexa scan, they have a z-score of -2 or less if they are under 50 years old, or a t-score of -2.5 or less if they are 50 years old or above. Only based on the criteria mentioned, does the person in the following dexa report have osteoporosis? Only consider the current status. Be as concise as possible. |

**Table 1. Prompts given to the models**
| Task | Prompt |
| --- | --- |
| Identifying dilated CBD | A normal common bile duct diameter is 6 mm and below for people aged 60 and under, and for each year after 60, 0.1 mm is added to the normal threshold. Only based on the criteria mentioned, does the person in the following ultrasound report have a dilated common bile duct? Only consider the current status. Be as concise as possible. |
| CBD: common bile duct; CT: computed tomography; PET: positron emission tomography |  |

### Data

We used two data sources for our study: our institution’s radiology reports database (for lung CT, PET, and DEXA data, use of data approved by the IRB, approval number 17-22317) and the MIMIC-III v1.4 dataset (19) (for ultrasound data). From each report type, we extracted 300 reports. These were selected either randomly (for MIMIC data) or consecutively (for institutional data) to reflect real-world performance. A sample size of 300 is suitable for an expected accuracy of 75% with a 95% level of confidence and an absolute margin of error of 5% (or an expected accuracy of 50% and an absolute margin of error of 6%). We believe the large sample size compensates to some degree for the lack of LLM response consistency experiments (repeat querying the same case) in our study, and allows for a comprehensive error analysis due to the variation between cases. For judgment tasks, the impression sections of the reports were removed. Further information on the data could be found in the supplementary methods.

### Ground truth and evaluation

Ground truths were extracted manually from the radiology reports. For evaluation, we solely considered the answer to the task, disregarding other potentially incorrect details or reasoning provided by the model. For example, if a task needed the size of a lung nodule and the response identified the size of a lung nodule correctly but misidentified its location, it was considered a correct response. Responses truncated due to token limits were only considered incorrect and incomplete if they could not be evaluated similarly to complete responses. For example, if the model stated “Yes” at the beginning of its response but ran out of tokens when explaining it, the initial “Yes” was evaluated. Further description of the evaluation method is presented in the supplementary methods.

We hypothesized that removing reports without the target values would change the results. Hence, we reported the results for the subset of reports that did contain target values as Non-NR (non-not reported) results.

### Error analysis

We reviewed all incorrect responses, identified error patterns, and summarized the findings quantitatively.

### Secondary experiment: answer-only output formatting

A common approach used with text generation models is to provide them with specific output formats. This not only might lead to more accurate results, but it could also provide outputs that are more comprehensible and can possibly be passed onto another automated system. Therefore, we performed the same experiments using formatting prompts. In this subtask, however, we used a stricter evaluation metric: a correct response contained only the correct output without any additional descriptive text. Mathematical signs such as the < sign were ignored in evaluation.

### Models

The non-reasoning large language model we used was the Llama 3.1 8B instruct version (20), and the reasoning models (21) we utilized were DeepSeek R1 distilled Llama 8b (22) (pseudo-reasoning and chain-of-thought version of Llama), OpenAI o1-mini, and OpenAI GPT-5-mini (low reasoning effort) (both true reasoning). Llama and DeepSeek models were accessed through the Hugging Face (23) transformers (24) package, and we queried our institution’s HIPAA-compliant Azure-based OpenAI models using the API. Hugging Face models had 16-bit optimization. We used a 256-token limit for the Llama model and a 1024-token limit for the reasoning models, as their reasoning process is included in their output tokens. The justifications behind setting token limits were two-fold: to account for resource constraints in the project that are also present in the real world, and to prevent scenarios of models entering repetitive or infinite response loops. Each report was included in a separate input to the model, consisting of the prompt and the report text.

### Statistical analyses

Agreement between two reviewers is demonstrated with Cohen’s κ. Model results are provided as proportional metrics: accuracy, sensitivity, and specificity. Confidence intervals were calculated using Wilson score interval (25) with continuity correction. Comparison between the original and formatted results and also between models in a task was done using McNemar’s test (via Python’s statsmodels package). Comparison between non-NR and NR cases and also between sensitivity and specificity in judgment tasks was done using Fisher’s exact test (via Python’s scipy package).

## Results

### Reviewer agreement

There was substantial agreement between reviewers in all tasks, with Cohen’s κ of 0.994 (5/900 disagreements) for extraction ground truths, 1.0 (0/900 disagreements) for judgment ground truths, 0.969 (6/3600 disagreements) for extraction results, 0.951 (26/3600 disagreements) for judgment results, 0.949 (10/3600 disagreements) for extraction error categorization, 0.880 (66/3600 disagreements) for judgment error categorization, and 0.991 (8/3600 disagreements) for extraction with formatting results.

### Number extraction tasks

Results for the extraction tasks can be found in Table 2 and Figure 1. Llama, DeepSeek, and o1-mini models performed best in extracting minimum T-score values (accuracy = 98.7% (96.4-99.6), 98.7% (96.4-99.6), and 99.3% (97.3-99.9), respectively), while the best performance of GPT-5-mini was in CBD diameter and nodule tasks (accuracy = 99.7% (97.9-100.0)). The worst performance for Llama and DeepSeek was in maximum CBD diameter extraction (accuracy = 86.0% (81.4-89.6) and 96.3% (93.3-98.1), respectively), for o1-mini was in maximum lung nodule size extraction (accuracy = 97.3% (94.6-98.8)), and for GPT-5-mini was the minimum T-score extraction task (accuracy = 99.0% (96.9-99.7)). Performances of Llama and o1-mini were significantly worse in NR cases of the maximum lung nodule diameter extraction task (p < 0.01). Llama had a statistically worse performance compared to all models in the CBD task.

**Table 2.** Extraction results.

|  | T-score |  | CBD diameter |  | Maximum lung nodule size |  |
| --- | --- | --- | --- | --- | --- | --- |
|  | All | Non-NR<br>(295,<br>98.3%) | All | Non-NR<br>(259,<br>86.3%) | All | Non-NR<br>(296,<br>98.7%) |
| **Llama 3.1 8b** | 98.7%<br>(96.4-99.6) | 98.6%<br>(96.3-99.6) | 86.0%<br>(81.4-89.6) | 84.6%<br>(79.4-88.6) | 95.3% (92.1-97.3) | 95.9%<br>(92.8-97.8)* |
| **DeepSeek R1**<br>**Distilled Llama 3.1**<br>**8b** | 98.7%<br>(96.4-99.6) | 98.6%<br>(96.3-99.6) | 96.3%<br>(93.3-98.1) | 96.9%<br>(93.8-98.6) | 97.7% (95.0-99.0) | 97.6%<br>(95.0-99.0) |
| **OpenAI o1-mini** | 99.3%<br>(97.3-99.9) | 99.3%<br>(97.3-99.9) | 99.0%<br>(96.9-99.7) | 99.2%<br>(96.9-99.9) | 97.3% (94.6-98.8) | 98.0%<br>(95.4-99.2)* |
| **OpenAI GPT-5-**<br>**mini** | 99.0%<br>(96.9-99.7) | 99.0%<br>(96.8-99.7) | 99.7%<br>(97.9-100.0) | 100.0%<br>(98.2-100.0) | 99.7% (97.9-100.0) | 99.7%<br>(97.8-100.0) |
| CBD: common bile duct, Non-NR: non-not reported, cases that had the target value; *: p < 0.05 between Non-NR and NR using Fisher's exact test |  |  |  |  |  |  |

**Figure 1.**
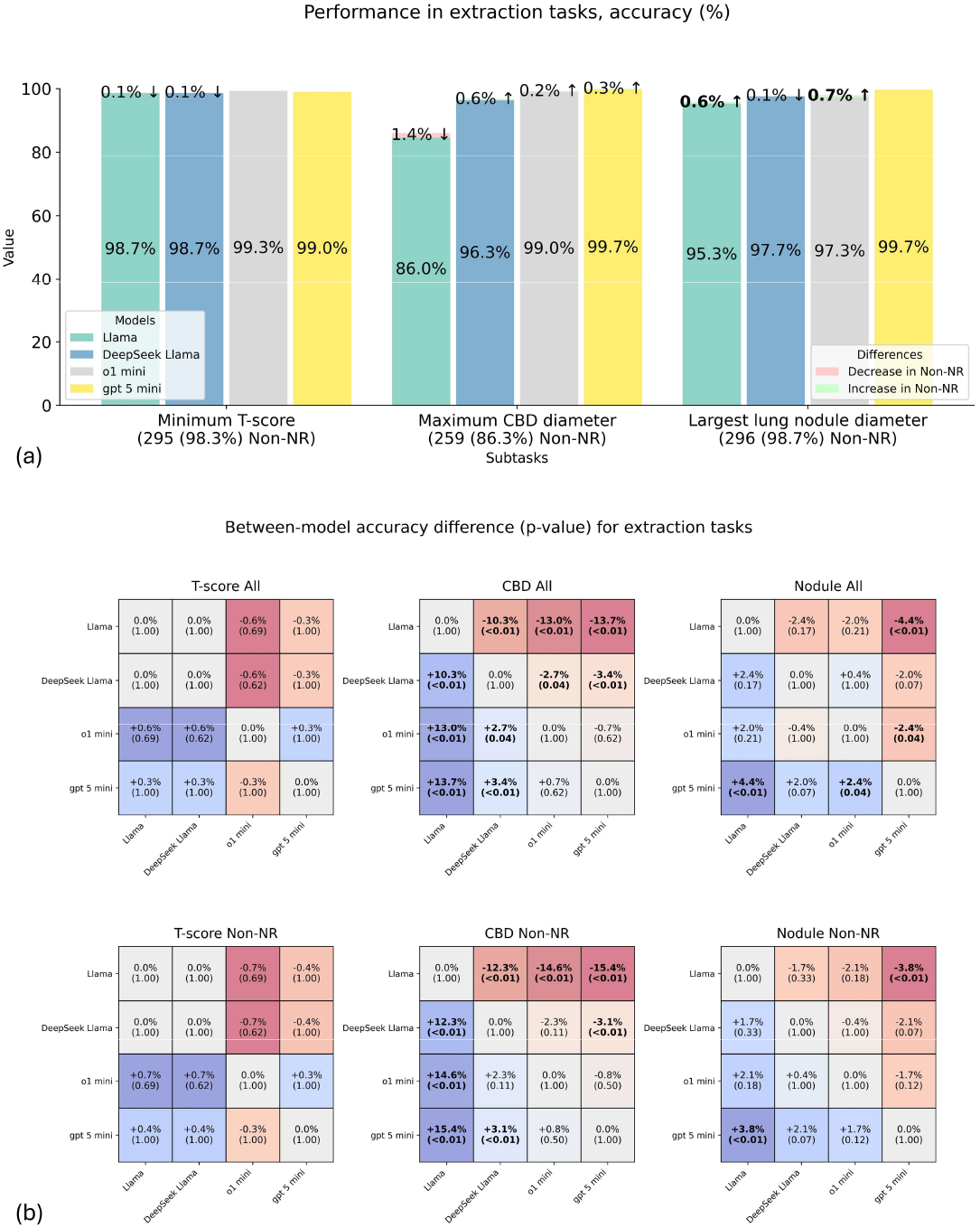
a) Performance in extraction tasks; b) between-model accuracy differences (p-values), bolded values indicate p-value < 0.05; CBD: common bile duct, Non-NR: non-not reported, cases that had the target value. The numbers on the bars are values for all cases. Bolded values indicate a statistically significant difference between Non-NR and NR cases using Fisher’s exact test (p < 0.05).

### Judgment tasks

Results for judgment tasks can be found in Table 3 and Figure 2. Accuracy of Llama ranged from 62.0% (56.2-67.5) to 96.0% (92.9-97.8), DeepSeek from 91.7% (87.8-94.4) to 96.7% (93.8-98.3), o1-mini from 91.7% (87.8-94.4) to 100.0% (98.4-100.0), and GPT-5-mini from 99.0% (96.9-99.7) to 100.0% (98.4-100.0). Llama had significantly lower accuracy than all models in PET and osteoporosis tasks, while GPT-5-mini had higher accuracy than all models in the PET task.

**Table 3.**
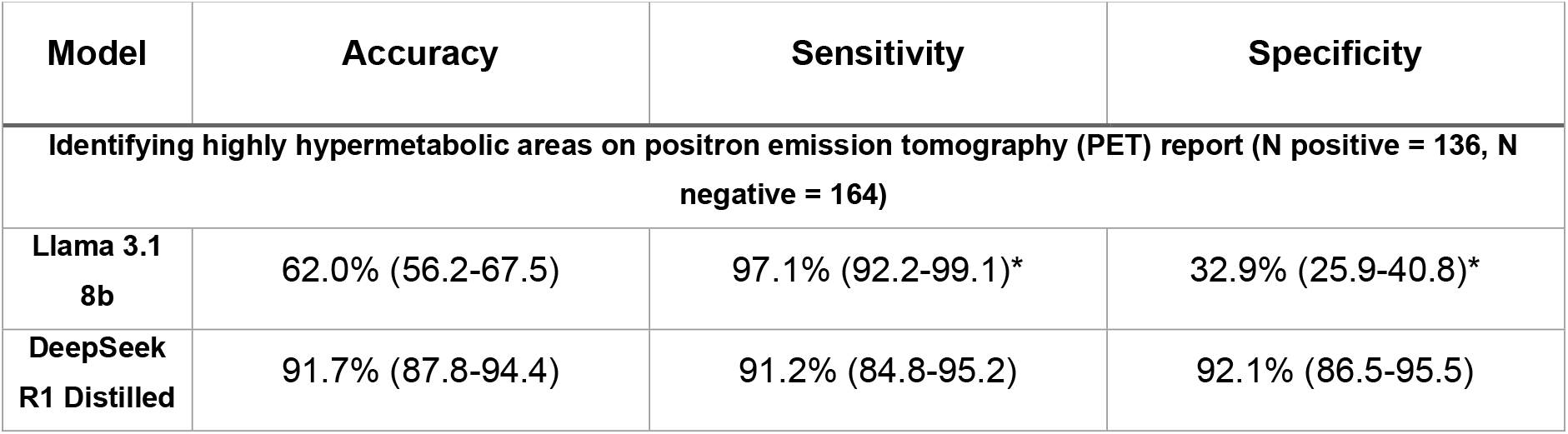
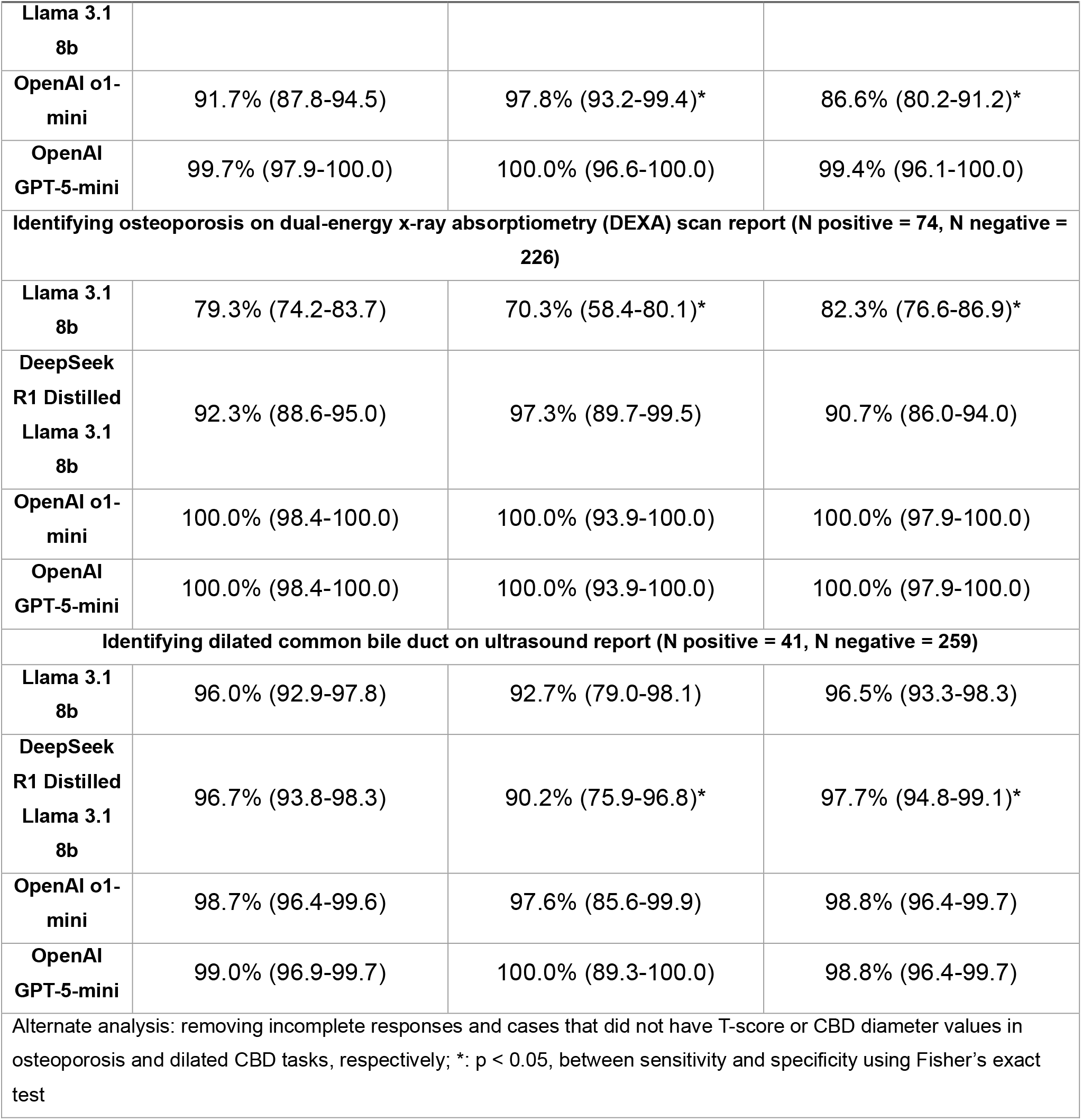
Judgment results.

| Model | Accuracy | Sensitivity | Specificity |
| --- | --- | --- | --- |
| Identifying highly hypermetabolic areas on positron emission tomography (PET) report (N positive = 136, N negative = 164) |  |  |  |
| **Llama 3.1**<br>**8b** | 62.0% (56.2-67.5) | 97.1% (92.2-99.1)* | 32.9% (25.9-40.8)* |
| **DeepSeek**<br>**R1 Distilled** | 91.7% (87.8-94.4) | 91.2% (84.8-95.2) | 92.1% (86.5-95.5) |

| Table 3. Judgment results |  |  |  |
| --- | --- | --- | --- |
| Model | Accuracy | Sensitivity | Specificity |
| Llama 3.1 8b |  |  |  |
| OpenAI o1-mini | 91.7% (87.8-94.5) | 97.8% (93.2-99.4)* | 86.6% (80.2-91.2)* |
| OpenAI GPT-5-mini | 99.7% (97.9-100.0) | 100.0% (96.6-100.0) | 99.4% (96.1-100.0) |
| Identifying osteoporosis on dual-energy x-ray absorptiometry (DEXA) scan report (N positive = 74, N negative = 226) |  |  |  |
| Llama 3.1 8b | 79.3% (74.2-83.7) | 70.3% (58.4-80.1)* | 82.3% (76.6-86.9)* |
| DeepSeek R1 Distilled Llama 3.1 8b | 92.3% (88.6-95.0) | 97.3% (89.7-99.5) | 90.7% (86.0-94.0) |
| OpenAI o1-mini | 100.0% (98.4-100.0) | 100.0% (93.9-100.0) | 100.0% (97.9-100.0) |
| OpenAI GPT-5-mini | 100.0% (98.4-100.0) | 100.0% (93.9-100.0) | 100.0% (97.9-100.0) |
| Identifying dilated common bile duct on ultrasound report (N positive = 41, N negative = 259) |  |  |  |
| Llama 3.1 8b | 96.0% (92.9-97.8) | 92.7% (79.0-98.1) | 96.5% (93.3-98.3) |
| DeepSeek R1 Distilled Llama 3.1 8b | 96.7% (93.8-98.3) | 90.2% (75.9-96.8)* | 97.7% (94.8-99.1)* |
| OpenAI o1-mini | 98.7% (96.4-99.6) | 97.6% (85.6-99.9) | 98.8% (96.4-99.7) |
| OpenAI GPT-5-mini | 99.0% (96.9-99.7) | 100.0% (89.3-100.0) | 98.8% (96.4-99.7) |
| Alternate analysis: removing incomplete responses and cases that did not have T-score or CBD diameter values in osteoporosis and dilated CBD tasks, respectively; *: $p < 0.05$, between sensitivity and specificity using Fisher's exact test | | | |

**Figure 2.**
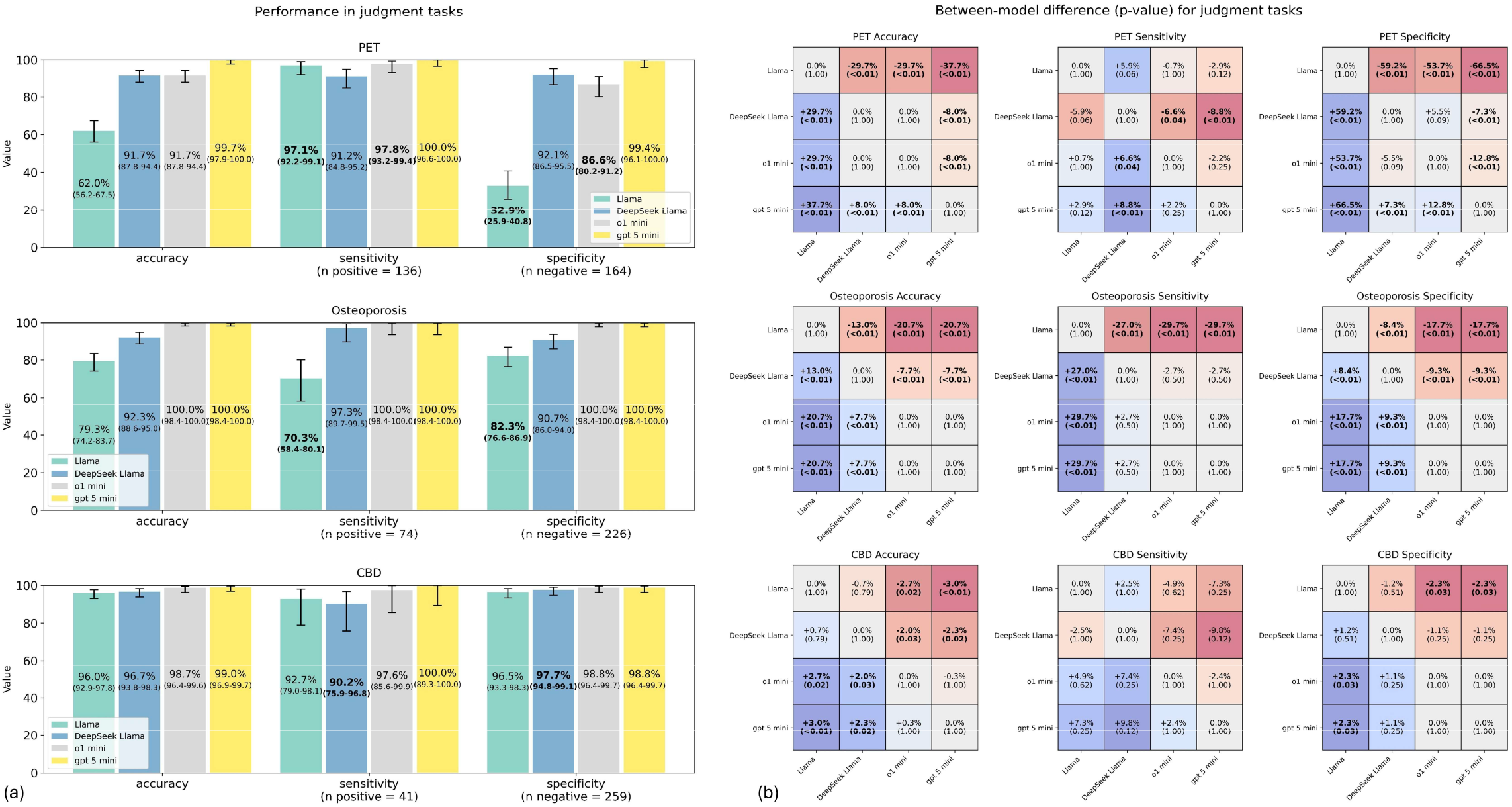
a) Performance in judgment tasks, bolded values indicate p-value < 0.05 between sensitivity and specificity using Fisher’s exact test; b) between-model accuracy differences (p-values), bolded values indicate p-value < 0.05; CBD: common bile duct.

### Error analysis

The results of the error analysis can be found in Figures 3 and 4. Overall, the most common error in a single task and a single model in terms of absolute number was assuming non-existent SUVMax values in the highly hypermetabolic area detection task by the Llama model (75 cases). GPT-5 and o1-mini models did not make any apparent mathematical errors, such as incorrect comparisons or calculations. Errors concerning medical concepts, such as understanding the differences between different biliary ducts and the notation of series/images in radiology reports, were seen among the outputs of all models. Error examples are presented in Supplementary Table 1.

**Figure 3.**
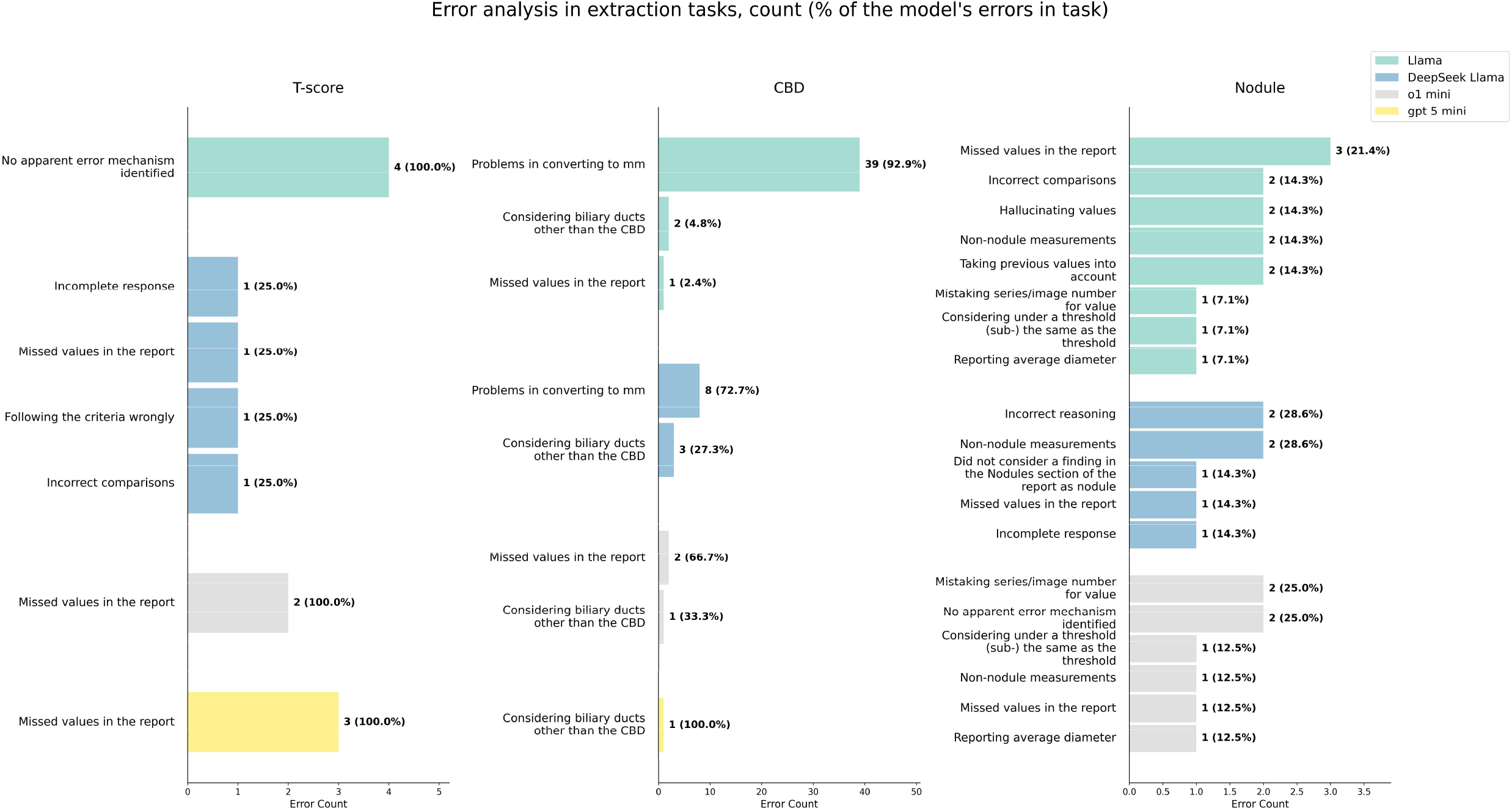
Error analysis in extraction tasks; CBD: common bile duct

**Figure 4.**
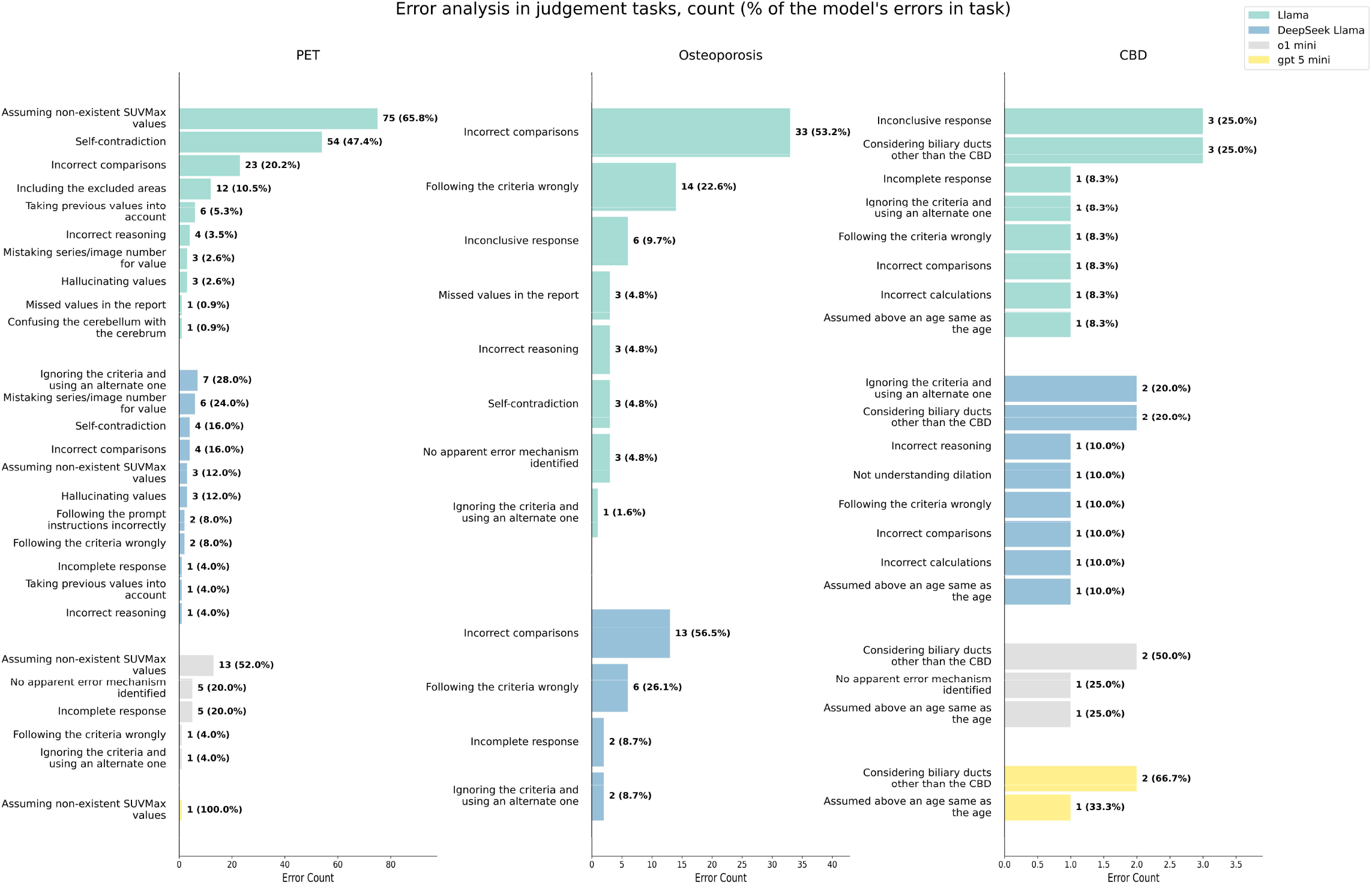
Error analysis in judgment tasks; CBD: common bile duct, PET: positron emission tomography. Percentages for each model might add up to more than 100% since some incorrect responses might have included multiple errors.

### Secondary experiment

Results for formatted output requests are presented in Table 4 and Figure 5. While Llama and DeepSeek models showed significantly decreased performance in almost all tasks (finding maximum nodule size was an exception for Llama), o1-mini and GPT-5-mini’s performance remained the same in all tasks, with minimum accuracies of 97.7% (95.0-99.0) (for maximum lung nodule size extraction) and 99.0% (96.9-99.7) (for minimum T-score extraction), respectively. Llama’s accuracy in Non-NR cases was significantly lower in minimum T-score extraction and higher in the other two tasks, and GPT-5-mini’s accuracy was significantly higher in Non-NR cases in the CBD diameter task. Llama was statistically the lowest-performing model in all tasks except the Non-NR CBD diameter extraction, where it had similar performance to DeepSeek Llama. GPT-5-mini was significantly superior to all models in the maximum nodule diameter extraction task.

**Table 4.**
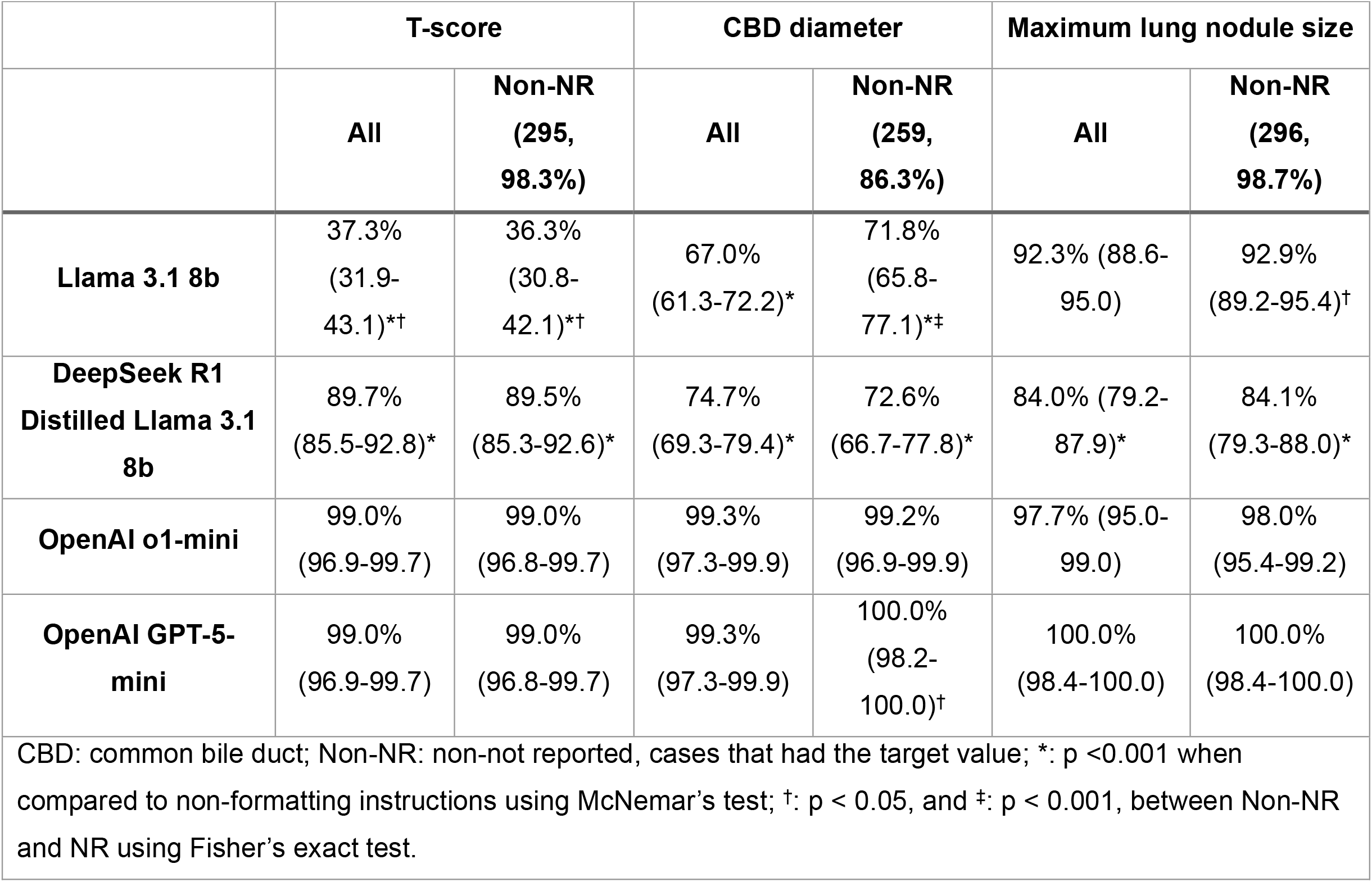
Extraction performance after formatting instructions.

**Figure 5.**
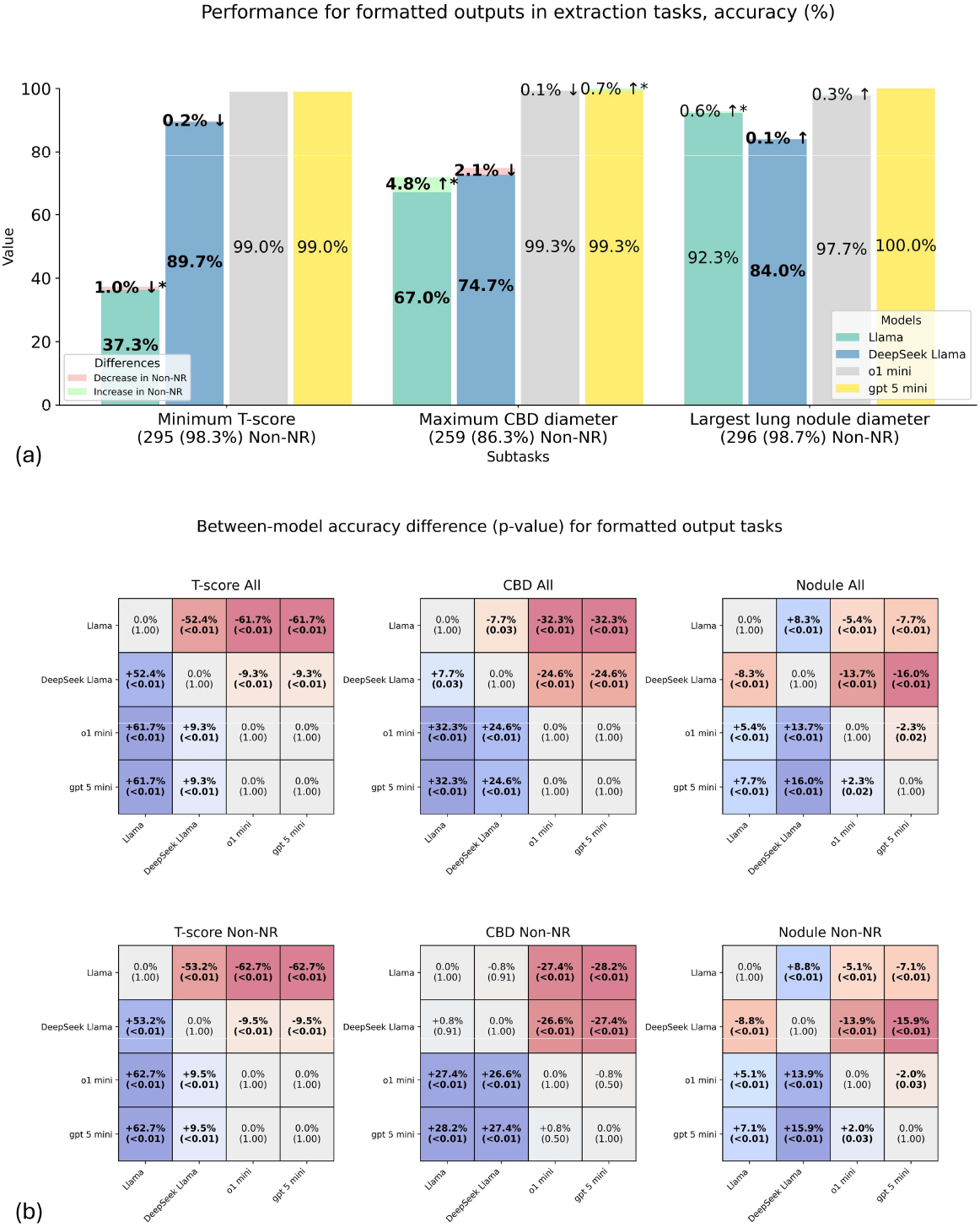
a) Performance for formatted outputs in extraction tasks; b) between-model accuracy differences (p-values), bolded values indicate p-value < 0.05; CBD: common bile duct, Non-NR: non-not reported, cases that had the target value. *: p < 0.05 for Non-NR vs NR cases using Fisher’s exact test. Bolded values indicate a statistically significant change compared to the non-formatted extraction task (p-value < 0.001) using McNemar’s test.

## Discussion

Our findings showed that the performance of LLMs in medical numerical tasks varied by model and task. As expected, true reasoning (i.e., GPT-5-mini and o1-mini) and pseudo-reasoning (i.e., DeepSeek) models performed better than Llama, the baseline LLM, in most of the tasks. Nonetheless, Llama 3.1 performed generally well on extraction tasks (+85% accuracy in extracting maximum CBD diameter and +95% in extracting maximum lung nodule diameter and minimum T-score). GPT-5-mini demonstrated highly robust performance with accuracies ≥99% in all tasks despite being a smaller version model and on low reasoning effort. Absence of the target value in the report mainly affected Llama, the non-reasoning model.

Some studies have shown that limiting the inputs to ones that include abnormal results generally improves LLM performance (1). Our results support these findings to some degree, although not consistently. Sensitivity was significantly higher than specificity in the PET judgment task for Llama and o1-mini, while the reverse was true for Llama in the osteoporosis task and DeepSeek Llama in the CBD task. Furthermore, in extraction tasks, the performance differed between reports with and without the target value in only a few instances, with a majority of them showing higher accuracies in cases with values (i.e., Non-NR cases). It is noteworthy that most of these significant differences were observed in experiments involving Llama. Hence, whether the model performs better in positive or negative cases depends on the model and the task, although this difference might be trivial in the case of newer models such as GPT-5-mini.

While most error types were shared between all models, mathematical errors were not seen in true reasoning models. Most errors that were also seen in more advanced models were related to medical knowledge to some degree, such as mistaking series numbers for desired values or other biliary ducts for the common bile duct. Hence, it may be possible that many true reasoning models achieve perfect results with well-optimized and task-specific prompting with adequate prerequisite information (such as medical concepts). It must be noted, however, that prompt engineering is a precise task that requires careful consideration. There are many caveats when designing the instructions passed to the model (26–28), and the most intuitive approaches might not lead to the best results.

It is a common practice to provide LLMs with requested output formats. In our study, instructing the model to provide the outputs in an answer-only format led to a significant deterioration of performance in non- and pseudo-reasoning models. This is in line with another study that demonstrated lower performance when specific output formats were requested, especially in reasoning-intensive tasks (29). Performance change varied between Llama and DeepSeek Llama depending on the task; nonetheless, DeepSeek Llama’s formatted performance was significantly lower than non-formatted instructions in all three tasks, suggesting that chain-of-thought reasoning does not necessarily lead to higher performance. o1-mini and GPT-5-mini, on the other hand, performed as well as the non-formatted stage with accuracies above 97%.

Our findings suggest the following recommendations when utilizing LLMs in radiological (and, to an extent, medical) tasks that involve numerical values or operations: 1) depending on the degree of acceptable error, smaller non-reasoning or pseudo-reasoning open-source models might provide adequate results, 2) this is less likely to be true when outputs are required to be in a specific format, such as pipeline extractions. Non-true-reasoning model performance drops significantly in such scenarios, and true reasoning models are especially recommended. 3) novel true reasoning models, even their smaller (mini) versions with low reasoning effort, which require fewer resources, perform with very high accuracy, and may obviate the need for more expensive larger models. 4) High accuracy is achievable with routine non-rigorous prompting and little prompt engineering; 5) medical concepts such as medical terms or medical text formats appear to be a pitfall of even more advanced LLMs, and providing these items as context in the prompts may lead to higher accuracies.

A few studies have previously addressed LLM capabilities in other medical tasks. Levin and colleagues (15) showed that LLMs could perform medication dosing calculations with high accuracy. A preprint (16) used a modified GPT-3 model on one thousand healthcare calculation tasks, achieving accuracies >80%. Furthermore, a recent analysis (30) demonstrated that removing lab values from medical scenarios hinders the diagnostic capabilities of LLMs, suggesting that LLMs understand and make use of the numerical values in medical scenarios. In a study involving MDCalc-derived (31) scenarios (7), Llama 3.1 and GPT 4 performance increased by multiple folds using methods such as retrieval-augmented generation (RAG) and utilizing task-specific tools. On the other hand, another study (13) experimented with chain-of-thought prompting on GPT 3.5 and found that it does not change the model’s performance on USMLE-style calculation tasks, remaining just under 80% accuracy.

There are some limitations to our study. We did not perform consistency analysis by running the prompt multiple times; however, we attempted to mitigate this issue by choosing a relatively large sample size of 300 for each task. The open-weight models we used were the smaller versions of their family, and using larger models may lead to better performance. Another limitation is that we set restrictions for the maximum output tokens due to time and resource constraints. Finally, although two reviewers evaluated the responses in our study, the task is to some degree subjective, and the results may slightly vary between different evaluators.

## Conclusion

True reasoning models, even their smaller versions, perform consistently well in radiological numerical tasks and are unlikely to make mathematical mistakes. While simpler models might achieve relatively high performance in simpler radiological numerical tasks, they are more prone to errors in more complex tasks, such as judgment tasks or tasks that require specific output formats.

## Data Availability

All data produced in the present study are available upon reasonable request to the authors

