## Supplementary for "Large language models in radiologic numerical tasks: A thorough evaluation and error analysis"

### Supplementary methods

#### Further information on data extraction

- For lung CT data, we used lung cancer screening CTs with the keyword “nodule” and optimized the results using the search engine’s built-in tool to maximize positive findings.
- For PET scans, we used the terms “index lesion” and “suvmax” in our search to extract the most relevant reports.

#### Considerations when evaluating responses

- In the extraction tasks, if there were multiple values in a report (e.g., both a range and an exact number), all of them were considered correct.
- If there were conflicting data in the reports, all were considered correct.
- When the correct answer was a range for a minimum/maximum task, both the range and one of its thresholds (depending on whether it was a maximum or minimum task) were considered correct. For instance, if the answer is 6-7 mm for finding the maximum CBD size, correct answers include 6-7 mm and 7 mm. However, if the range specifically mentioned that the measurements are below or above a certain value, that value was not considered correct (e.g., nodules measuring <5 mm).
- For the common bile duct tasks, while we did not accept common hepatic duct measurements if explicitly stated, we accepted “common duct” measurements.
- In the formatted output tasks, we accepted values without the < sign, as the prompt asked for solely the value.
- In the CBD and osteoporosis judgment tasks, if there was not enough data to make the judgment (e.g., no CBD or age values), both “cannot determine” and negative answers were considered correct. The case was treated as a negative case for analysis.

**Supplementary Table S1. Error examples**

| **Model** | Task | Error category | Specific error | Relevant report sections | Relevant sections of the model output |
|---|---|---|---|---|---|
| Llama | Extracting minimum T-score | No apparent error mechanism | | RESULTS:<br>L1-L4 Total<br>BMD values= 0.859 T-score= -1.7 Z-score= 0.1<br><br>Total Femur<br>BMD values= 0.714 T-score= -1.9 Z-score= -0.6<br>Femur Neck<br>BMD values= 0.632 T-score= -2.0 Z-score= -0.4 | -1.7 |
| | Extracting maximum CBD diameter | Not following the instructions | Incorrect conversion to mm | The common bile duct is normal measuring 0.5 cm. | 0.5 cm |
| | Extracting maximum CBD diameter | Not understanding medical concepts | Confusing other biliary ducts with the common bile duct | The common hepatic duct is dilated at the porta hepatis measuring approximately 15 mm | The maximum common bile duct diameter is approximately 15 mm. |
| | Extracting maximum CBD diameter | Reading the report incorrectly | Missing a value | There is marked intra- and extrahepatic biliary dilatation, with the intrahepatic ducts measuring up to 6 mm and common duct measuring up to 1 cm | 6mm. |
| | Extracting maximum lung nodule size | Not following the instructions | Considering the previous lung nodule dimension | The previously described 3 mm nodule in the left upper lobe has since resolved | The largest reported lung nodule size is 3 mm. |
| | Extracting maximum lung nodule size | Not following the instructions | Reporting the average diameters instead of the maximum diameter | 1.5 cm (1.6 x 1.3 cm groundglass nodule left upper lobe | The largest reported lung nodule size is 1.5 cm. |

| | | | | | |
|---|---|---|---|---|---|
| | Extracting maximum lung nodule size | Reading the report incorrectly | Missing a value | Scattered 3 to 4 mm solid pulmonary nodules including the right apex | No nodule size reported. |
| | Extracting maximum lung nodule size | Reading the report incorrectly | Not a lung nodule | LUNG NODULES:<br>Unchanged 4 mm nodule along the right minor fissure (series 2 image 101) disease.<br>LUNGS (OTHER FINDINGS):<br>The lungs are otherwise clear.<br>PLEURA:<br>Normal.<br>MEDIASTINUM:<br>Unchanged soft tissue nodule in the anterior mediastinum measuring 9 mm in short axis. | The largest reported lung nodule size is 9 mm. |
| | Extracting maximum lung nodule size | Reading the report incorrectly | Hallucinating a value | LUNG NODULES:<br>- Lateral basal left lower lobe solid noncalcified nodule (image 357) measuring 4 mm<br>- Lingular groundglass nodule (series 3, image 273) measuring 1.3 x 0.8 cm<br>- Left apical nodule (image 91) measuring 2 mm | The largest reported lung nodule size is 10 mm, which is the size of the lingular groundglass nodule. |
| | Extracting maximum lung nodule size | Incorrect mathematical operations | Incorrect comparisons | Right upper lobe mixed solid and groundglass nodule (series 3 image 56) overall measuring 2.9 x 1.6 cm | The largest reported lung nodule size is 12 mm. |
| | Extracting maximum lung nodule size | Not understanding medical concepts | Mistaking the series/image number for measurement | LUNG NODULES:<br>Scattered pulmonary micronodules including a pulmonary nodule within the right lower lobe (3/202). | The largest reported lung nodule size is 3 mm. |
| | Extracting maximum lung nodule size | Reading the report incorrectly/Incorrect reasoning | Considering under a threshold (sub-) the same as the threshold | Multiple scattered sub-6 mm solid pulmonary nodules | 6 mm. |
| | Identifying highly hypermetabolic areas on PET | Reading the report incorrectly | Hallucinating an SUVMax value | Increased size and erosive changes of the hypermetabolic left lateral | Based on the given criteria (SUVMax > 5), the highly hypermetabolic areas in |

| | | | | |
|---|---|---|---|---|
| | | | 10th rib measuring 2.3 x 1.3 cm (series 401 image 225), previously 1.8 x 1.1 cm. | the PET report, excluding the cerebellum and pituitary, are:<br><br>Left lateral 10th rib (SUV max 6.4, but only 4.3 is mentioned, however, 6.4 is the value in the report for the series 401 image 225) |
| Identifying highly hypermetabolic areas on PET | Not following the instructions | Assuming non-existent SUVMax values | Spleen: Measures 8.5 cm in craniocaudal dimension. No hypermetabolism above background.<br><br>Abdomen/Pelvis: Similar size of right adnexal cystic lesion measuring 5.1 x 6.9 cm (series 201 image 184), previously 5.0 x 7.1 cm on most recent prior exam and 2.8 x 4.5 cm on PET/CT 5/23/2022. Myomatous uterus. | Based on the given criteria (SUVMax > 5, excluding cerebellum and pituitary), the current highly hypermetabolic areas are:<br><br>1. Right adnexal cystic lesion (SUVMax not specified, but size is 5.1 x 6.9 cm)<br>2. Myomatous uterus (SUVMax not specified)<br>3. Spleen (SUVMax not specified, but measures 8.5 cm in craniocaudal dimension)<br><br>Note that the SUVMax values are not provided for the spleen and the adnexal cystic lesion, but they are mentioned as having some level of hypermetabolism. The myomatous uterus is also mentioned as having hypermetabolism, but no SUVMax value is provided. |
| Identifying highly hypermetabolic areas on PET | Incorrect mathematical operations | Incorrect comparisons | Nodular uptake in the left erector spinae musculature with SUV max 4.3<br><br>Left proximal femoral diaphysis: Status post curettage and placement of a left intramedullary nail with low-level patchy tracer uptake in the surrounding musculature with SUV max 4.1 | Based on the given criteria (SUVMax > 5), the highly hypermetabolic areas in the PET report, excluding the cerebellum and pituitary, are:<br><br>Left erector spinae musculature (SUV max 4.3)<br><br>Left proximal femoral diaphysis surrounding musculature (SUV max 4.1) |
| Identifying highly hypermetabolic areas on PET | Reading the report incorrectly | Missing a value | Hypermetabolic rounded lymph node anterior to the distal gastric body greater | Based on the provided information, there are no highly hypermetabolic areas (SUVMax > 5) in the current PET |

| | | | | |
|---|---|---|---|---|
| | | | curvature measuring 8 mm short axis, SUV max 6.2 | report, excluding the cerebellum and pituitary. |
| Identifying highly hypermetabolic areas on PET | Logical and language errors | Incorrect reasoning | | Based on the provided PET report, the highly hypermetabolic areas with SUVMax greater than 5, excluding the cerebellum and pituitary, are not mentioned. |
| Identifying highly hypermetabolic areas on PET | Logical and language errors | Self-contradiction | Small focus of hypermetabolism along the inferior endplate of L4 (series 3 image 226, SUV Max 4.7) | Based on the given criteria (SUVMax > 5, excluding cerebellum and pituitary), there is one highly hypermetabolic area mentioned in the report:<br><br>- Inferior endplate of L4 (SUV Max 4.7) is not a highly hypermetabolic area as it is less than 5. |
| Identifying highly hypermetabolic areas on PET | Not following the instructions | Not excluding the cerebellum and the pituitary | | Based on the given criteria (SUVMax greater than 5, excluding cerebellum and pituitary), the only highly hypermetabolic area in this PET report is the cerebellum with a SUVMax of 7.2. |
| Identifying highly hypermetabolic areas on PET | Not following the instructions | Considering the previous SUVMax value | Centrally necrotic right level 2/3 lymph nodes: Decreased in size. Previously 4.3 x 2.8 cm. Decreased hypermetabolism now with patchy hypermetabolism, SUV Max 2.5, previously SUV Max 16.3 | Centrally necrotic right level 2/3 lymph nodes: SUV Max 2.5 is below the criteria, but the prior SUV Max was 16.3. |
| Identifying highly hypermetabolic areas on PET | Reading the report incorrectly | Mistaking the series number for SUVMax | …for example a left common iliac lymph node measures 0.6 cm (201/180)<br><br>Complex, heterogeneous, predominantly osteoblastic lesion in the left anterior acetabular column with the highest level of hypermetabolism at the periphery of left acetabulum (201/204) | Based on the SUVMax values, the highly hypermetabolic areas (SUVMax > 5) in the current PET report, excluding the cerebellum and pituitary, are:<br><br>1. The periphery of the left acetabulum (SUVMax: 201/204)<br>2. A left common iliac lymph node measures 0.6 cm (SUVMax: 201/180) |

| | | | | |
|---|---|---|---|---|
| Identifying highly hypermetabolic areas on PET | Not understanding medical concepts | Confusing cerebrum with cerebellum | Cerebellar SUVmax is 9.6. | Cerebral SUVmax is 9.6 (not specified which part, but assuming it's the brain) |
| Determining the presence of osteoporosis | Incorrect mathematical operations | Incorrect comparisons | 6X years old<br><br>L1-L3 Total<br>BMD values= 0.897 T-score= -1.1 Z-score= 0.8<br><br>Total Femur<br>BMD values= 0.695 T-score= -2.0 Z-score= -0.7<br>Femur Neck<br>BMD values= 0.585 T-score= -2.4 Z-score= -0.8 | Yes, the person has osteoporosis based on the current dexa report. The T-score of -2.4 in the Femur Neck is less than -2.5, meeting the criteria for osteoporosis in a 6X -year-old. |
| Determining the presence of osteoporosis | Reading the report incorrectly | Missing a value | L1-L4 Total<br>BMD values= 0.611 T-score= -4.0 Z-score= -1.7<br><br>Total Femur<br>BMD values= 0.678 T-score= -2.2 Z-score= -0.6<br>Femur Neck<br>BMD values= 0.582 T-score= -2.4 Z-score= -0.5 | Based on the criteria, the person does not have osteoporosis in the total femur or femur neck areas since their T-scores are -2.2 and -2.4, respectively, which are above -2.5. |
| Determining the presence of osteoporosis | Logical and language errors | No conclusive response | 7X years old<br><br>L1-L4 Total<br>BMD values= 0.934 T-score= -1.0 Z-score= 1.5<br><br>Total Femur<br>BMD values= 0.758 T-score= -1.5 Z-score= 0.4<br>Femur Neck<br>BMD values= 0.637 T-score= -1.9 Z-score= 0.3 | Based on the given dexa report, the person has a T-score of -1.0 in the lumbar spine, which is not -2.5 or less, and a T-score of -1.5 in the total femur, and a T-score of -1.9 in the femur neck, which are also not -2.5 or less.<br><br>However, the person is 7X years old, which is 50 years or above. For the total femur, the T-score is -1.5, and for the femur neck, the T-score is -1.9. |
| Determining the presence of osteoporosis | Logical and language errors | Self-contradiction | Distal radius<br>One third (33%) region of interest (ROI) | According to the criteria, a T-score of -2.5 or less is required for a diagnosis of osteoporosis. Since the person's T-scores are all less than -2.5, they do not |

| | | | | |
|---|---|---|---|---|
| | | | BMD values=0.668 T-score=-2.8 Z-score=-1.8<br>Total ROI<br>BMD values=0.544 T-score=-2.7 Z-score=-1.8<br><br>Ultradistal ROI<br>BMD values=0.376 T-score=-2.9 Z-score=-1.8 | meet the criteria for osteoporosis based on the T-score. |
| Determining the presence of osteoporosis | Logical and language errors | Incorrect reasoning | [Patient in their 70s]<br><br>L1-L4 Total<br>BMD values= 0.844 T-score= -1.8 Z-score= 0.5<br><br>Total Femur<br>BMD values= 0.716 T-score= -1.8 Z-score= -0.2<br>Femur Neck<br>BMD values= 0.510 T-score= -3.1 Z-score= -1.1 | No, the person does not have osteoporosis according to the provided criteria.<br><br>The criteria for osteoporosis in a 50-year-old or above is a t-score of -2.5 or less. The t-scores provided for the lumbar spine and total femur are -1.8, which does not meet this criteria. The t-score for the femur neck is -3.1, but this is the only area that meets the criteria for osteoporosis, however, this value is for the femur neck, not the lumbar spine or total femur which are the areas evaluated in the report. |
| Determining the presence of osteoporosis | Not following the instructions | Ignoring the provided criteria and using an alternate one | [Patient in their 10s]<br><br>L1 - L4 Total<br>BMD values= 0.781 T-score= Z-score= -2.4<br>Only if > 10 years:<br>Total Femur<br>BMD values= 0.737 T-score= Z-score= -2.2 | No, the person does not have osteoporosis based on the given criteria. The criteria for osteoporosis in children under 50 years old is a z-score of -2 or less, but in this report, the z-score is -2.8, which is less than -2. However, the WHO criteria of osteoporosis do not apply for children, which means the criteria provided in the question do not apply to this case. |
| Determining the presence of osteoporosis | Not following the instructions | Following the criteria incorrectly | 9X years old<br><br>L1-L3 Total<br>BMD values= 0.579 T-score= -4.0 Z-score= -1.2<br><br>Total Femur | Based on the provided criteria, the person does not have osteoporosis at any evaluated area.<br><br>At L1-L3, the T-score is -4.0, which is less than -2.5, but the person is 9X years old, so we use the Z-score of -1.2 to evaluate osteoporosis. Since the Z- |

| | | | | |
|---|---|---|---|---|
| | | | BMD values= 0.582 T-score= -3.0 Z-score= -0.6<br>Femur Neck<br>BMD values= 0.418 T-score= -3.9 Z-score= -1.4 | score is not -2 or less, this area does not indicate osteoporosis.<br><br>At the Total Femur, the T-score is -3.0, which is less than -2.5, but the person is 9X years old, so we use the Z-score of -0.6 to evaluate osteoporosis. Since the Z-score is not -2 or less, this area does not indicate osteoporosis.<br><br>At the Femur Neck, the T-score is -3.9, which is less than -2.5, but the person is 9X years old, so we use the Z-score of -1.4 to evaluate osteoporosis. Since the Z-score is not -2 or less, this area does not indicate osteoporosis. |
| Determining the presence of osteoporosis | No apparent error mechanism | | 8X years old<br><br>L1-L4 Total<br>BMD values= 1.142 T-score= 0.9 Z-score= 3.6<br><br>Total Femur<br>BMD values= 0.820 T-score= -1.0 Z-score= 1.1<br>Femur Neck<br>BMD values= 0.613 T-score= -2.1 Z-score= 0.2<br>Positioning, artifacts and morphologic findings: None. | The person has osteoporosis based on the T-score of -2.1 or less in the Femur Neck, which is below 50 years of age is not a consideration, and the person is 8X years old, which makes them post-menopausal and older than 50 years. |
| Identifying dilated CBD | Incorrect mathematical operations | Wrong calculations | 6X -year-old female with right upper quadrant pain status post cholecystectomy | Since the person is 6X years old, we add 0.1 mm for each year after 60. X years after 60 would be 6X. So, the normal threshold would be 6 mm + (X * 0.1 mm) = Y mm. |
| Identifying dilated CBD | Incorrect mathematical operations | Wrong comparisons | [Patient in their 30s] | No, the person does not have a dilated common bile duct. The diameter of 7 mm is below the threshold for a person [in their 30s], which would be 6 mm or below. |

| | | | | | |
|---|---|---|---|---|---|
| | Identifying dilated CBD | Not following the instructions | Following the criteria incorrectly | 4X year old man<br>the common duct measures 0.3 cm. | The person is 4X years old, so we need to add 0.1 mm for each year after 60. 4X - 60 = -1Y years (since 4X is less than 60), so we need to subtract 1.Y * 0.1 mm = 0.1Y mm from the normal threshold.<br>Normal threshold = 6 mm - 0.1Y mm = 5.Z mm |
| | Identifying dilated CBD | Not following the instructions | Ignoring the provided criteria and using an alternate one | 6X -year-old female<br>The common bile duct is slightly bigger than the prior study of measuring 7 mm | No, the person does not have a dilated common bile duct. The common bile duct measures 7 mm, which is the upper limit of normal for her age (6X years old). The normal threshold is 6 mm for people 60 and under, and 0.1 mm is added for each year after 60. Therefore, the threshold for a 6X -year-old person is 6 + (6X - 60) * 0.1 = 6.X mm. |
| | Identifying dilated CBD | Logical and language errors | No conclusive response | 8X year old man<br>The common bile duct measures 3 mm. | The person is 8X years old, so the threshold is 6.3 mm.<br>The common bile duct measures 3 mm, which is below the threshold. |
| | Identifying dilated CBD | Logical and language errors | Assuming above an age same as the age | [**Age over 9X **] | The person is 9X years old. |
| | Identifying dilated CBD | Not understanding medical concepts | Confusing other biliary ducts with the common bile duct | The common hepatic duct remains prominent at 7 mm | Based on the report, the common hepatic duct diameter is 7 mm, which is not within the normal threshold for a 6X -year-old man (normal threshold would be 6 mm + 0.1 mm * (6X -60) = 6.X mm). Therefore, the person has a dilated common bile duct. |
| DeepSeek | Extracting minimum T-score | Reading the report incorrectly | Missing a value | L1-L4 Total<br>BMD values= 1.029 T-score= -0.6 Z-score= 0.5<br><br>Total Femur<br>BMD values= 0.864 T-score= -1.1 Z-score= -0.3<br>Femur Neck | <think><br>Looking at the report, the results section has two T-scores: one for L1-L4 Total and another for the Total Femur. Double-checking the report, I don't see any other T-scores mentioned, so I'm confident that -1.1 is the correct extraction.<br></think> |

| | | | | |
|---|---|---|---|---|
| | | | BMD values= 0.761 T-score= -1.2 Z-score= 0.1 | -1.1 |
| Extracting minimum T-score | Not following the instructions | Following the criteria incorrectly | Z-score = -2.8<br>T-score= Z-score= -2.4<br>T-score= Z-score= -2.2 | The current minimum T-score is **-2.8**. |
| Extracting minimum T-score | Incorrect mathematical operations | Incorrect comparisons | L2-L4 Total<br>BMD values= 0.859 T-score= -3.0 Z-score= -1.2<br><br>Total Femur<br>BMD values= 0.759 T-score= -1.7 Z-score= -0.9<br>Femur Neck<br>BMD values= 0.678 T-score= -1.9 Z-score= -0.8 | <think><br>The L2-L4 Total has a T-score of -3.0, which is quite low. The Total Femur is -1.7, and the Femur Neck is -1.9. Since the user wants the minimum T-score, I should pick the smallest number among these. The Femur Neck has the lowest T-score at -1.9.<br></think><br><br>The current minimum T-score is **-1.9**. |
| Extracting maximum CBD diameter | Not understanding medical concepts | Confusing other biliary ducts with the common bile duct | The common hepatic duct is dilated at the porta hepatis measuring approximately 15 mm | <think><br>Then, in the final report, under findings, it says, "The common hepatic duct is dilated at the porta hepatis measuring approximately 15 mm, there is no dilation of the intra-hepatic ducts."<br><br>So, the maximum common bile duct diameter mentioned is 15 mm. The distal common bile duct isn't visualized, but the main measurement given is 15 mm at the porta hepatis.<br></think><br><br>The maximum common bile duct diameter reported is **15 mm**. |
| Extracting maximum CBD diameter | Not following the instructions | Incorrect conversion to mm | The extrahepatic common duct is at the upper limits of normal measuring 0.6 cm. | The maximum common bile duct diameter reported in the ultrasound is 0.6 cm.<br><br>0.6 cm |

| | | | | |
|---|---|---|---|---|
| Extracting maximum lung nodule size | Not following the instructions | Not a lung nodule | LUNG NODULES:<br>Unchanged 4 mm nodule along the right minor fissure (series 2 image 101) disease.<br><br>MEDIASTINUM:<br>Unchanged soft tissue nodule in the anterior mediastinum measuring 9 mm in short axis. | <think><br>The findings mention two nodules: one is a 4 mm nodule unchanged, and another is a 9 mm soft tissue nodule in the anterior mediastinum.<br><br>So, between these two, the 9 mm is larger than the 4 mm. The user is asking for the largest, so 9 mm is the one.<br></think><br><br>The largest reported lung nodule size is **9 mm**. |
| Extracting maximum lung nodule size | Reading the report incorrectly | Did not take into account a finding in the Nodules section of the report | LUNG NODULES:<br>Similar right apical scarring with an area of slightly increased density (3/63) measuring up to 5 mm. | no nodule size reported |
| Extracting maximum lung nodule size | Reading the report incorrectly | Missing a value | the largest a right upper lobe nodule measuring 5 mm | <think><br>I need to focus only on the current nodules and ignore any previous sizes. The largest mentioned in the current findings are both 4 mm. There's no mention of any larger nodules in the current report. So, the largest reported size is 4 mm.<br></think><br><br>The largest reported lung nodule size from the current findings is 4 mm.<br><br>**Answer:** 4 mm |
| Extracting maximum lung nodule size | Logical and language errors | Incorrect reasoning | Previously noted 5 mm right upper lobe nodule and 4 mm left lower lobe nodule are unchanged. | <think><br>Previously noted were a 5 mm right upper lobe nodule and a 4 mm left lower lobe nodule, which are unchanged.<br><br>So, the current nodules mentioned are numerous, but their sizes aren't specified. The only sizes given are the |

| | | | | |
|---|---|---|---|---|
| | | | | previous ones, which are 5 mm and 4 mm.<br></think><br><br>no nodule size reported |
| Identifying highly hypermetabolic areas on PET | Reading the report incorrectly | Hallucinating SUVMax values | Distended appendix measuring up to 1.2 cm with associated hypermetabolism, compatible with ongoing appendicitis.<br><br>Hypermetabolism in distal thoracic esophagus, most commonly reflecting esophagitis. | <think><br>Other findings include the appendix with hypermetabolism, indicating possible appendicitis. There's also hypermetabolism in the distal thoracic esophagus, likely esophagitis. Both of these have SUVMax values above 5, so they fit the criteria.<br><br>I should list these areas: distended appendix and distal thoracic esophagus. The bones' hypermetabolism is noted but not included since it's nonspecific. I need to make sure I'm only including areas with SUVMax >5 and excluding cerebellum and pituitary.<br><br>So, the answer should mention these two areas without including the cerebellum or pituitary.<br></think><br><br>The PET report indicates hypermetabolism in the following areas:<br><br>- **Distended appendix**: Compatible with ongoing appendicitis.<br>- **Distal thoracic esophagus**: Likely reflecting esophagitis.<br><br>These areas meet the SUVMax >5 criterion. |
| Identifying highly hypermetabolic areas on PET | Reading the report incorrectly | Assuming non-existent SUVMax values | Hypermetabolism within the endometrium and left ovary is favored physiologic in a premenopausal woman. | <think><br>The report says "Hypermetabolism in the endometrium and left ovary." It doesn't provide exact SUVMax values, but since they're noted as |

| | | | | |
|---|---|---|---|---|
| | | | | hypermetabolic, and the threshold is 5, it's likely that their SUVMax is above 5. So these areas would qualify as highly hypermetabolic.<br></think><br><br>The PET report indicates hypermetabolism in the endometrium and left ovary, both of which are SUVMax greater than 5. These areas qualify as highly hypermetabolic. |
| Identifying highly hypermetabolic areas on PET | Incorrect mathematical operations | Incorrect comparisons | Increased size and erosive changes of the hypermetabolic left lateral 10th rib measuring 2.3 x 1.3 cm (series 401 image 225), previously 1.8 x 1.1 cm. Similar size and appearance of the lytic lesion in the posterior C2 arch with low-level FDG uptake. Nodular uptake in the left erector spinae musculature with SUV max 4.3 and without CT correlate, nonspecific. | <think><br>The left lateral 10th rib has SUVmax 4.3. That's above 5, so that's a hypermetabolic area.<br></think><br><br>The only highly hypermetabolic area identified, excluding the cerebellum and pituitary, is the left lateral 10th rib lesion with an SUVmax of 4.3. |
| Identifying highly hypermetabolic areas on PET | Not following the instructions | Considering the previous SUVMax value | Cardiophrenic node: 3.6 x 2.3 cm, SUV Max 30.3, (Se/Im 2/148) on prior, now resolved. | The only highly hypermetabolic area, besides the excluded cerebellum and pituitary, is the cardiophrenic node with an SUV Max of 30.3. |
| Identifying highly hypermetabolic areas on PET | Not following the instructions | Following the criteria incorrectly | New hypermetabolism within the 0.9 cm right external iliac node (series 201 image 195) with SUV max of 5.4 | Then there's a new hypermetabolism in the right external iliac node, 0.9 cm, SUVmax 5.4. Hmm, 5.4 is just below 5.5, so technically it's not over 5. Wait, no, 5.4 is less than 5.5, so it doesn't meet the criteria. So that's not a highly hypermetabolic area. |
| Identifying highly hypermetabolic areas on PET | Not following the instructions | Ignoring the provided criteria and using an alternate one | Midline abdominal wall mass originating from rectus abdominis muscle decreased size and hypermetabolism: Similar in size measuring | <think><br>Next, looking at the index lesions. The first one is a midline abdominal wall mass. Its SUVmax is 3.3, which is above 2.8 from before. |

| | | | | |
|---|---|---|---|---|
| | | | approximately 9.3 x 5.5 cm. SUV Max 3.3, previously SUV Max 2.8 | So, the key areas are the abdominal wall mass and possibly the spleen if its SUVmax is above 5.<br></think><br><br>The PET report indicates hypermetabolic areas as follows:<br><br>**Abdominal Wall Mass**: Demonstrates an SUVmax of 3.3, which is above the previous value of 2.8, indicating increased hypermetabolism. |
| Identifying highly hypermetabolic areas on PET | Not following the instructions | Following the prompt instructions incorrectly | Asymmetric low-level hypermetabolism of the right lower paraspinal and medial left greater than right upper thigh musculature with SUV max 5.9. Additional low-level hypermetabolism of the bilateral gluteal musculature with SUV max of 7.0. | <think><br>Moving on to the muscles, the right lower paraspinal area has an SUVMax of 5.9. That's just above 5, so it's borderline. Then the medial left upper thigh is 5.9 as well. The bilateral gluteal muscles have an SUVMax of 7.0, which is definitely above 5.<br><br>I need to check if these muscle areas are considered hypermetabolic. The report notes that these are low-level hypermetabolisms, which could be due to the patient's known myopathy. Myopathy often involves muscle-related issues, so the hypermetabolism in the muscles might be related to that rather than cancer.<br><br>Since the report says there's no evidence of hypermetabolic malignancy, the areas with SUVMax above 5 in the muscles aren't indicative of disease. Therefore, the only areas meeting the SUVMax >5 criteria are the muscles, but they're likely nonspecific and related to the patient's existing condition.<br><br>So, in conclusion, besides the excluded cerebellum and pituitary, there are the |

| | | | | |
|---|---|---|---|---|
| | | | | muscles mentioned. However, they don't meet the criteria for hypermetabolic malignancy, so they're probably not significant in this context.<br></think><br><br>No, there are no other highly hypermetabolic areas with SUVMax greater than 5 in the current PET report, excluding the cerebellum and pituitary. |
| Identifying highly hypermetabolic areas on PET | Not understanding medical concepts | Mistaking the series/image number for SUVMax | Hypermetabolic cutaneous lesions involving the left lower cheek and left temporal scalp, similar compared to prior (3775/458 and 417). | <think><br>Now, the report talks about hypermetabolic cutaneous lesions on the left lower cheek and left temporal scalp. The SUV values are 3775 and 417, which are both way above 5. So these areas are highly hypermetabolic.<br></think><br><br>The highly hypermetabolic areas identified in the current PET/CT report, excluding the cerebellum and pituitary, include:<br><br>**Left lower cheek and left temporal scalp**: SUV values 3775 and 417. |
| Identifying highly hypermetabolic areas on PET | Logical and language errors | Incorrect reasoning | T5 vertebral body lytic lesion: More sclerotic in appearance, measuring 1.8 x 1.1 cm, SUV Max 5.3 | <think><br>T5 vertebral body: SUVMax is 5.3. That's just above 5, but not over 5, so it doesn't qualify as highly hypermetabolic.<br><br>So, after reviewing all the lesions, the only area that meets the SUVMax >5 is the cerebellum, which we're excluding. All other areas have SUVMax 5.3 or lower in the current exam. Therefore, there are no other highly hypermetabolic areas beyond the cerebellum and pituitary in this report.<br></think> |

| | | | | | |
|---|---|---|---|---|---|
| | | | | | No, there are no other highly hypermetabolic areas in the PET report with an SUVMax greater than 5, excluding the cerebellum and pituitary. |
| | Identifying highly hypermetabolic areas on PET | Logical and language errors | Self-contradiction | A hypermetabolic irregular right upper lobe nodule: 2.2 x 2.6 cm cm, SUV Max 11.1 | <think><br>First, I'll go through the findings section. It mentions a hypermetabolic right upper lobe nodule with an SUVMax of 11.1.<br><br>So, putting it all together, the only highly hypermetabolic area in the current report, apart from cerebellum and pituitary, is the right upper lobe nodule. I should make sure to exclude the cerebellum and pituitary as per the user's request. Therefore, the answer is just that one nodule.<br></think><br><br>No other highly hypermetabolic areas with SUVMax greater than 5 are identified in the current PET report. |
| | Determining the presence of osteoporosis | Incorrect mathematical operations | Incorrect comparisons | [Patient in their 70s]<br><br>L1, L3-L4 Total<br>BMD values= 0.893 T-score= -1.5 Z-score= 0.9<br><br>Total Femur<br>BMD values= 0.895 T-score= -0.5 Z-score= 1.2<br>Femur Neck<br>BMD values= 0.609 T-score= -2.2 Z-score= -0.3 | <think><br>Since the T-score is -2.2, which is less than -2.5, this meets the osteoporosis criterion for someone over 50.<br></think><br><br>Based on the criteria, the individual meets the diagnosis of osteoporosis due to the T-score of -2.2 in the Femur Neck, which is ≤-2.5 for someone aged 50 or older.<br><br>**Answer:** Yes, the person has osteoporosis. |
| | Determining the presence of osteoporosis | Not following the instructions | Following the criteria incorrectly | [Patient in their 40s]<br><br>L1-L4 Total<br>BMD values= 0.998 T-score= -0.4 Z-score= 0.2 | <think><br>Then there's the Femur Neck with a BMD of 0.612, T-score of -2.1, and Z-score of -1.5. The T-score here is -2.1, which is way below the -1.0 cutoff for |

| | | | | |
|---|---|---|---|---|
| | | | Total Femur<br>BMD values= 0.828 T-score= -0.9 Z-score= -0.5<br>Femur Neck<br>BMD values= 0.612 T-score= -2.1 Z-score= -1.5 | osteoporosis in postmenopausal women.<br></think><br><br>Based on the dexa report, the individual meets the osteoporosis criteria due to a T-score of -2.1 in the femur neck, which is below the -2 cutoff for someone under 50.<br><br>**Answer:** The person has osteoporosis. |
| Determining the presence of osteoporosis | Not following the instructions | Ignoring the provided criteria and using an alternate one | | <think><br>So, putting it all together, despite not knowing the exact age and bone density scores, the severe vertebral fractures in a person with osteopenia meet the criteria for osteoporosis.<br></think><br><br>Based on the provided DXA report, the individual has multiple severe compression fractures (Grade 3) in the thoracolumbar spine, along with a history of osteopenia. These findings, particularly the severe fractures in a person with osteopenia, strongly suggest a diagnosis of osteoporosis. The absence of explicit bone density scores does not negate the significance of the fractures, which are indicative of the condition. Therefore, the individual meets the criteria for osteoporosis.<br><br>**Answer:** Yes, the person has osteoporosis. |
| Identifying dilated CBD | Not understanding medical concepts | Confusing other biliary ducts with the common bile duct | The common hepatic duct remains prominent at 7 mm | <think><br>Looking at the report, the common hepatic duct is measured at 7 mm.<br></think><br><br>The 6X -year-old's normal common bile duct threshold is 6.X mm. The |

| | | | | |
|---|---|---|---|---|
| | | | | ultrasound reports a 7 mm measurement, which is above this threshold, indicating dilatation.<br><br>**Answer:** Yes, the common bile duct is dilated, measuring 7 mm in a 6X - year-old, exceeding the normal threshold of 6.X mm. |
| Identifying dilated CBD | Not following the instructions | Ignoring the provided criteria and using an alternate one | [Patient in their 40s]<br><br>The common duct measures 6.6 mm. | <think><br>I think in this case, since the person is under 60, the normal is 6 mm. 6.6 is only a 0.6 increase, which might not be enough to be considered dilated based on the given criteria. The report also says there's no dilation elsewhere, so it's just this one measurement. So probably, he doesn't have a dilated common bile duct based on the current status and the given criteria.<br></think><br><br>Based on the ultrasound report and the provided criteria, the individual does not have a dilated common bile duct. The common bile duct measures 6.6 mm, which is slightly above the normal threshold of 6 mm for someone under 60. However, there is no indication of intra- or extra-hepatic duct dilation, and the criteria do not consider a 0.6 mm increase as dilated. Therefore, the current status does not meet the criteria for dilation.<br><br>**Answer:** No, the person does not have a dilated common bile duct based on the provided criteria. |
| Identifying dilated CBD | Not following the instructions | Following the criteria incorrectly | [Patient in their 50s]<br><br>the common bile duct measures approximately 0.6 cm. | <think><br>At 60 years, it's 6 mm. So for each year before 60, we subtract 0.1 mm.<br></think> |

| | | | | |
|---|---|---|---|---|
| | | | | The person is 5X years old. The normal common bile duct diameter threshold is calculated as follows:<br><br>- At 60 years: 6 mm<br>- For each year before 60, subtract 0.1 mm: X years * 0.1 mm = 0.X mm<br>- Normal threshold = 6 mm - 0.X mm = 5.X mm<br><br>The ultrasound report states the common bile duct measures 0.6 cm, which is 6 mm. Since 6 mm > 5.X mm, it is considered dilated.<br><br>**Answer:** Yes, the person has a dilated common bile duct. |
| Identifying dilated CBD | Incorrect mathematical operations | Incorrect comparisons | [Patient in their 50s]<br><br>the common duct measures 0.5 cm | <think><br>I think I got it. The common bile duct is 5 mm, which is above the normal threshold of 6 mm<br></think><br><br>The common bile duct measures 5 mm, which is above the normal threshold of 6 mm for a 5X -year-old, indicating dilatation.<br><br>**Answer:** Yes, the common bile duct is dilated. |
| Identifying dilated CBD | Incorrect mathematical operations | Wrong calculations | 6X -year-old female<br><br>common bile duct is slightly bigger than the prior study of measuring 7 mm | <think><br>So, at 6X, that would be 6 + 0.1* X = 7.X mm.<br></think><br><br>Based on the provided information, the common bile duct diameter is X mm for a 6X -year-old patient, which is within the normal threshold of 7.X mm. Therefore, the common bile duct is not considered dilated under the given criteria. |

| | | | | |
|---|---|---|---|---|
| | | | | **Answer:** No, the common bile duct is not dilated. |
| Identifying dilated CBD | Logical and language errors | Incorrect understanding of the word dilated | 6XF<br><br>The common duct measures 2 mm. | <think><br>So, putting it all together, the CBD is 2 mm, which is less than the normal threshold of 6.X mm for a 6X -year-old. Therefore, it's considered dilated.<br></think><br><br>The common bile duct measures 2 mm, which is below the normal threshold for a 6X -year-old (6.X mm). Therefore, it is considered dilated.<br><br>**Answer:** Yes, the person has a dilated common bile duct. |
| Identifying dilated CBD | Logical and language errors | Assuming above an age same as the age | [**Age over 90 **] | <think><br>Let's see, from 60 to 90 is 30 years. So, 30 years times 0.1 mm per year equals 3 mm added.<br></think><br>The person is over 90, so her normal CBD diameter threshold is 9 mm. The report states the CBD measures 1.4 cm (14 mm), which is dilated.<br><br>**Answer:** Yes, the common bile duct is dilated. |
| Identifying dilated CBD | Logical and language errors | Incorrect reasoning | 4X year old woman<br><br>Prominent common bile duct measuring up to 8 mm | <think><br>Since she's 4X, she's Y years under 60. Each year after 60 adds 0.1 mm, so Y years would add Y * 0.1 = Z mm. Therefore, her normal threshold is 6 + Z = T mm.<br></think><br><br>Based on the provided criteria, the 4X -year-old woman's common bile duct diameter of T mm is within the normal range for her age (4X + Y years after 60, adding Z mm to the normal 6 mm, |

| | | | | | |
|---|---|---|---|---|---|
| | | | | | totaling T mm). Thus, she does not have a dilated common bile duct.<br><br>**Answer:** No, the person does not have a dilated common bile duct. |
| o1-mini | Extracting minimum T-score | Reading the report incorrectly | Missing a value | L1 - L4 Total<br>BMD values= 0.748 T-score=<br>Z-score= -2.5<br>Only if > 10 years:<br>Total Femur<br>BMD values= 0.771 T-score=<br>Z-score= -1.7 | not reported |
| | Extracting maximum CBD diameter | Not understanding medical concepts | Confusing other biliary ducts with the common bile duct | The common hepatic duct is dilated at the porta hepatis measuring approximately 15 mm | 15 |
| | Extracting maximum CBD diameter | Reading the report incorrectly | Missing a value | There is no intrahepatic biliary ductal dilation, though the common bile duct is again prominent, measuring 1.1 cm, as seen on recent CTs | not reported |
| | Extracting maximum lung nodule size | Reading the report incorrectly/Incorrect reasoning | Considering under a threshold (sub-) the same as the threshold | A few scattered sub-4 mm subpleural nodules. | 4 mm |
| | Extracting maximum lung nodule size | Reading the report incorrectly | Missing a value | Previously noted 5 mm right upper lobe nodule (series/image 2/56) and 4 mm left lower lobe nodule (3/337) are unchanged. | no nodule size reported |
| | Extracting maximum lung nodule size | Not following the instructions | Not a lung nodule | LUNG NODULES:<br>Unchanged 4 mm nodule along the right minor fissure (series 2 image 101) disease.<br>MEDIASTINUM:<br>Unchanged soft tissue nodule in the anterior mediastinum measuring 9 mm in short axis. | 9 mm |

| | | | | |
|---|---|---|---|---|
| Extracting maximum lung nodule size | Not following the instructions | Reporting the average diameters instead of the maximum diameter | 1.5 cm (1.6 x 1.3 cm groundglass nodule left upper lobe | 1.5 cm |
| Extracting maximum lung nodule size | Not understanding medical concepts | Mistaking the series/image number for measurement | Pulmonary nodules within the right upper lobe are slightly larger and have developed a new rim of groundglass (3/222) | 3 mm |
| Extracting maximum lung nodule size | No apparent error mechanism | | LUNG NODULES:<br>Increase in size of region of irregular consolidation in the right upper lobe currently measuring 2.8 x 2.6 cm. A satellite nodules/region of consolidation has also increased measuring 10 mm, previously 5 mm.<br>New left upper lobe nodule along the major fissure currently measuring 5 mm (series 2 image 85). New 6 mm nodule in the periphery of the left upper lobe on series 2 image 130.<br>Stable 5 mm nodule/nodular scar at the left lung apex on series 2 image 53. | 10 mm |
| Identifying highly hypermetabolic areas on PET | Reading the report incorrectly | Assuming non-existent SUVMax values | Prostatomegaly with calcifications and increased nonspecific multifocal FDG uptake.<br><br>Increased nonspecific multifocal uptake in the prostate gland. Correlate clinically. | Yes, the current PET report identifies increased nonspecific multifocal uptake in the prostate gland with SUVMax greater than 5. |
| Identifying highly hypermetabolic areas on PET | Not following the instructions | Following the criteria incorrectly | Hypermetabolic left outer breast mass: Approximately 1.6 x 1.3 cm , SUV Max 5. | Yes, the dominant left outer breast mass with an SUVmax of 5. |

| | | | | | |
|---|---|---|---|---|---|
| | Identifying highly hypermetabolic areas on PET | Not following the instructions | Ignoring the provided criteria and using an alternate one | Focal hypermetabolism in the right lateral 10th rib with associated fracture deformity. | Yes, there is a highly hypermetabolic area:<br><br>- **Right lateral 10th rib** with associated fracture deformity. |
| | Determining the presence of osteoporosis | | | n/a | |
| | Identifying dilated CBD | Not understanding medical concepts | Confusing other biliary ducts with the common bile duct | 6X old man<br><br>The common hepatic duct remains prominent at 7 mm | Yes, the common hepatic duct is dilated. For a 6X -year-old, the normal threshold is 6.X mm (6 mm + X×0.1 mm). The reported diameter of 7 mm exceeds this threshold. |
| | Identifying dilated CBD | Logical and language errors | Assuming above an age same as the age | [**Age over 90 **] | Yes, the common bile duct is dilated.<br><br>**Calculation:**<br>- **Age:** 9X years<br>- **Normal threshold:** 6 mm + (9X - 60) × 0.1 mm = 9 mm<br>- **Measured CBD:** 14 mm<br><br>Since 14 mm > 9 mm, the common bile duct is dilated. |
| | Identifying dilated CBD | No apparent error mechanism | | 7X year old<br><br>The common bile duct is measuring 8 mm which is at the upper limits of normal for a patient of this age. | No, the common bile duct measuring 8 mm is within the normal range for a 7X -year-old. |
| GPT-5 mini | Extracting minimum T-score | Reading the report incorrectly | Missing a value | L1 - L4 Total<br>BMD values= 0.748 T-score=<br>Z-score= -2.5<br>Only if > 10 years:<br>Total Femur<br>BMD values= 0.771 T-score=<br>Z-score= -1.7 | not reported |

| | | | | |
|---|---|---|---|---|
| Extracting maximum CBD diameter | Not understanding medical concepts | Confusing other biliary ducts with the common bile duct | The common hepatic duct remains prominent at 7 mm | 7 mm |
| Extracting maximum lung nodule size | | n/a | | |
| Identifying highly hypermetabolic areas on PET | Not following the instructions | Assuming non-existent SUVMax values | New intense focus of hypermetabolism without clear correlate on nondiagnostic CT study, possibly located within the left masseter muscle or anterior left parotid gland.<br><br>new hypermetabolic focus without clear CT correlate which may be located in the left parotid gland or masseter muscle. | Yes — the new intense hypermetabolic focus in the left masseter/anterior left parotid region. No other current areas meet SUVmax >5 per the report. |
| Determining the presence of osteoporosis | | n/a | | |
| Identifying dilated CBD | Not understanding medical concepts | Confusing other biliary ducts with the common bile duct | 6X year old man<br><br>The common hepatic duct remains prominent at 7 mm | Age 6X → normal threshold = 6.0 + (6X −60)*0.1 = 6.X mm. Reported duct = 7 mm, which is >6.X mm. Conclusion: yes — the duct is mildly dilated. |
| Identifying dilated CBD | Logical and language errors | Assuming above an age same as the age | [**Age over 90 **] | Yes. At age ~90 the normal threshold ≈ 6 mm + (90−60)×0.1 mm = 9 mm (0.9 cm). The CBD measures 1.4 cm (14 mm), which exceeds that threshold, so it is dilated. |

CBD: common bile duct; PET: positron emission tomography; Age values in blue were modified to comply with medRxiv policy
